## Supplementary material for "Integration of clinical, pathological, radiological, and transcriptomic data improves the prediction of first-line immunotherapy outcome in metastatic non-small cell lung cancer": Annex

**Annex 1: Glossary of clinical features**

- **age:** Age of the patient at the diagnosis of the metastatic disease.
- **albumin:** Serum albumin (g/l).
- **alk:** Detected ALK mutation prior to the initiation of first-line therapy (1: detected, 0: otherwise).
- **asat:** Serum Aspartate Aminotransferase (AST) (IU/l).
- **bmi:** Body Mass Index (kg/m^2^) at the diagnosis of the metastatic disease.
- **braf:** Detected BRAF mutation prior to the initiation of first-line therapy (1: detected, 0: otherwise).
- **chemotherapy:** Whether the patient received chemotherapy+pembrolizumab or not (1: pembrolizumab+chemotherapy, 0: pembrolizumab alone).
- **ecog:** ECOG status (Eastern Cooperative Oncology Group) at the diagnosis of the metastatic disease, one-hot encoded into 4 binary features (ecog_0, ecog_1, ecog_2, ecog_4).
- **egfr:** Detected EGFR mutation prior to the initiation of first-line therapy (1: detected, 0: otherwise).
- **erbb2:** Detected ERBB2 mutation prior to the initiation of first-line therapy (1: detected, 0: otherwise).
- **height:** Height of the patient (cm).
- **histo:** Histology one-hot encoded into 4 binary features (histo_adeno for adenocarcinomas, histo_squamous for squamous cell carcinomas, histo_other-nsclc for other NSCLC subtypes, and histo_other for other unspecified subtypes).
- **kras:** Detected KRAS mutation prior to the initiation of first-line therapy (1: detected, 0: otherwise).
- **ldh:** Serum Lactate dehydrogenase (LDH) (IU/l).
- **lung_surgery:** Whether the patient had a lung surgery prior to the first-line therapy (1: prior lung surgery, 0: otherwise).
- **lymphocytes:** Circulating lymphocytes count (10e9/l).
- **meta_brain:** Whether a brain metastasis was detected prior to the initiation of first-line therapy (1: detected brain metastasis, 0: otherwise).
- **met:** Detected MET mutation prior to the initiation of first-line therapy (1: detected, 0: otherwise).
- **neutrophils:** Circulating neutrophils count (10e9/l).
- **neutrophils/lymphocytes:** Neutrophils-to-lymphocytes ration (NLR).
- **n_preceding_cancers**: Number of preceding cancers.
- **other_mutations:** Detected mutations (different from BRAF, EGFR, ERBB2, KRAS, MET, or ROS1) prior to the initiation of first-line therapy (1: detected, 0: otherwise)
- **pack_years:** Smoking history in pack-years (i.e., equivalent of smoking one pack of 20 cigarettes a day for one year).
- **pdl1:** PD-L1 status (1: positive detection of PD-L1 expression with immunohistochemistry prior to the initiation of first-line therapy, 0: otherwise). If several tests were performed before the first-line therapy a positive test prevails over negative tests.
- **pdl1_tps:** PD-L1 Tumor Proportion Score (TPS). If several tests were performed before the initiation of first-line therapy, the maximum score across the tests is chosen. For patient with negative PD-L1 status this feature is set to 0.
- **ros1:** Detected ROS1 mutation prior to the initiation of first-line therapy (1: detected, 0: otherwise).
- **sex:** Sex of the patient (0: man, 1: woman).
- **smoking:** Smoking status with 3 categories (i.e., 3 binary variables), smoking_never, smoking_past, and smoking_current.
- **tils:** Detection of Tumor-Infiltrating Lymphocytes (TILs) prior to the initiation of first-line therapy (1: positive detection, 0: otherwise).
- **weight:** Weight of the patient at the diagnosis of the metastatic disease (kg).

**Annex 2: Glossary of radiomics features**

- **Distance dispersion:** Quartile dispersion of the distances between each tumor region's centroid and the global centroid.
- **Dmax:** Largest distance between the centroids of two lesions normalized by the body surface area.
- **liver_SUVMean:** Mean of SUV values in a spherical ROI manually delineated in a healthy part of the liver on the PET scan.
- **Nb invaded organs:** Number of invaded organs visible on the PET scan, including the lungs, sub- and supra-diaphragmatic lymph nodes, the pleura, the liver, the bones, the adrenal gland, and a final category for other regions.
- **N1_Suvmax_max/mean/std:** Maximum/Mean/Standard deviation value of the SUVmax of the segmented lesions located in regions associated with the N1 stage (i.e., ipsilateral hilar and mediastinal-hilar lymph nodes). 0 if no lesions in these regions.
- **N1_TMTV:** Total Metabolic Tumor Volume computed with the segmented lesions located in regions associated with the N1 stage. 0 if no lesions in these regions.
- **N2_Suvmax_max/mean/std:** Maximum/Mean/Standard deviation value of the SUVmax of the segmented lesions located in regions associated with the N2 stage (i.e., ipsilateral mediastinal lymph nodes and subcarinal lymph nodes). 0 if no lesions in these regions.
- **N2_TMTV:** Total Metabolic Tumor Volume computed with the segmented lesions located in regions associated with the N2 stage. 0 if no lesions in these regions.
- **N3_Suvmax_max/mean/std:** Maximum/Mean/Standard deviation value of the SUVmax of the segmented lesions located in regions associated with the N3 stage (i.e., contralateral mediastinal and hilar lymph nodes and supraclavicular lymph nodes). 0 if no lesions in these regions.
- **N3_TMTV:** Total Metabolic Tumor Volume computed with the segmented lesions located in regions associated with the N3 stage. 0 if no lesions in these regions.
- **M1a_Suvmax_max/mean/std:** Maximum/Mean/Standard deviation value of the SUVmax of the segmented lesions located in regions associated with the M1a stage (i.e., contralateral lung metastases and pleural metastases). 0 if no lesions in these regions.
- **M1a_TMTV:** Total Metabolic Tumor Volume computed with the segmented lesions located in regions associated with the M1a stage (excluding diffuse pleural metastases). 0 if no lesions in these regions.
- **M1bc_Suvmax_max/mean/std:** Maximum/Mean/Standard deviation value of the SUVmax of the segmented lesions located in regions associated with the M1b and M1c stages (i.e., extrathoracic metastases). 0 if no lesions in these regions.
- **M1bc_TMTV:** Total Metabolic Tumor Volume computed with the segmented lesions located in regions associated with the M1b and M1c stages (excluding diffuse subdiaphragmatic metastases). 0 if no lesions in these regions.
- **spleen_SUVMean:** Mean of SUV values in a spherical ROI manually delineated in a healthy part of the spleen on the PET scan.
- **TMTV:** Total Metabolic Tumor Volume computed with all the segmented lesions (excluding diffuse lesions corresponding to lymphangitic spread, diffuse pleural metastases, diffuse myocardial metastases, and diffuse subdiaphragmatic metastases).
- **T_Suvmax_max/mean/std:** Maximum/Mean/Standard deviation value of the SUVmax of the segmented lesions located in regions associated with the T stage (i.e., primary lung tumor and ipsilateral lung metastases). 0 if no lesions in these regions.
- **T_TMTV:** Total Metabolic Tumor Volume computed with the segmented lesions located in regions associated with the T stage (excluding lymphangitic spread). 0 if no lesions in these regions.
