## Supplementary figures and tables for "Integration of clinical, pathological, radiological, and transcriptomic data improves the prediction of first-line immunotherapy outcome in metastatic non-small cell lung cancer"

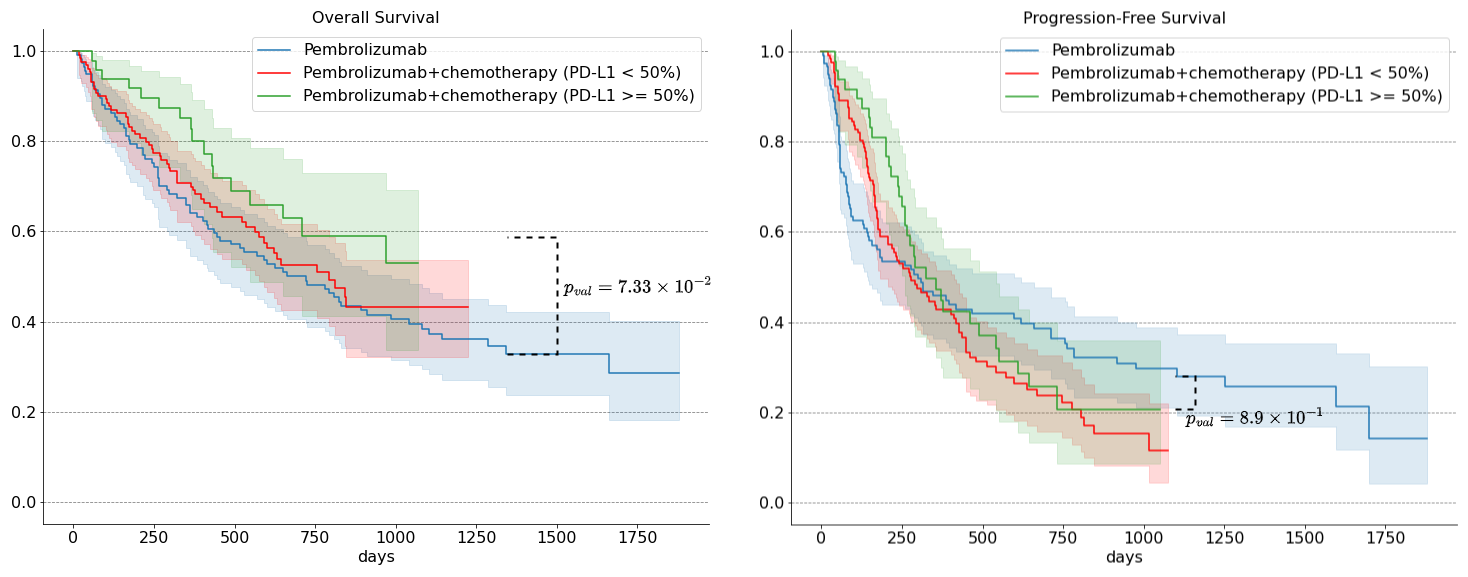
**Figure s1:** OS and PFS Kaplan-Meier survival curves for the whole NSCLC cohort (n=311 for OS and n=316 for PFS) with 95% confidence interval. Patients are first stratified with respect to their first-line therapy (pembrolizumab vs pembrolizumab + chemotherapy). Patients treated with pembrolizumab + chemotherapy are also stratified with respect to their PD-L1 expression (PD-L1 TPS < 50% vs PD-L1 TPS ≥ 50%). Log-rank p-values are reported to assess the separation of survival curves.

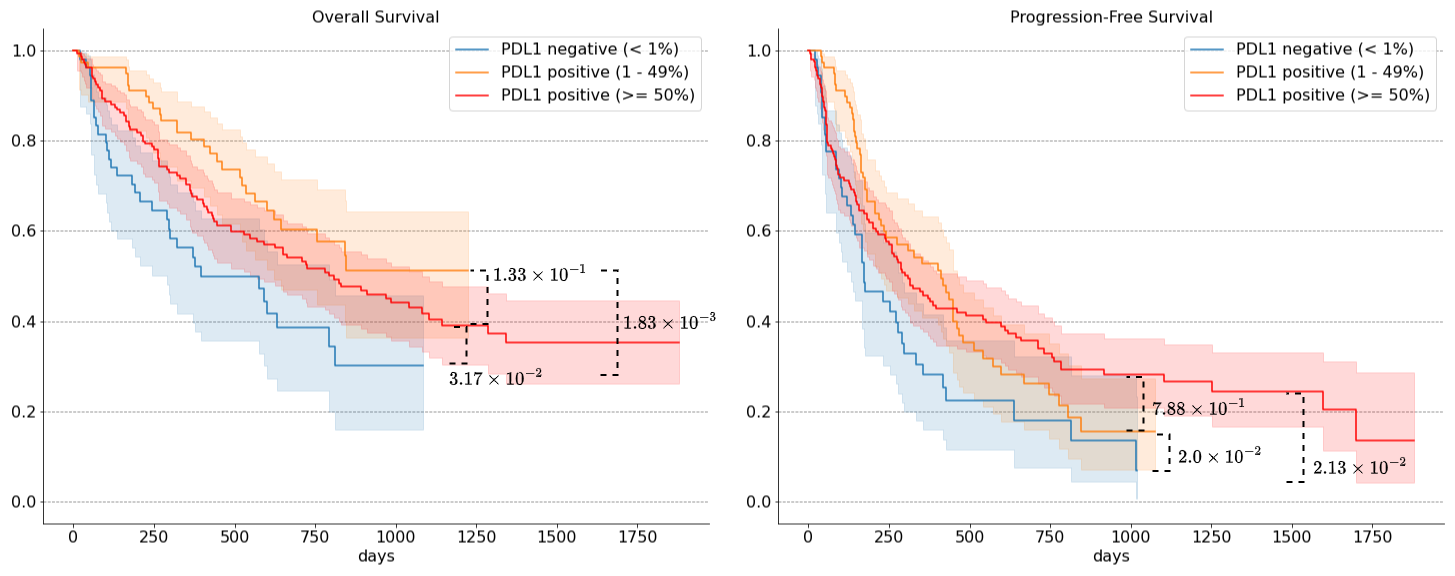

**Figure s2:** OS and PFS Kaplan-Meier survival curves for the patients with available PD-L1 expression (n=295 for OS and n=300 for PFS) with 95% confidence interval. Patients are first stratified with respect to PD-L1 expression (PD-L1 negative (i.e., TPS < 1%) vs PD-L1 positive with TPS < 50% vs PD-L1 positive with TPS ≥ 50%). Log-rank p-values are reported to assess the separation of the survival curves.

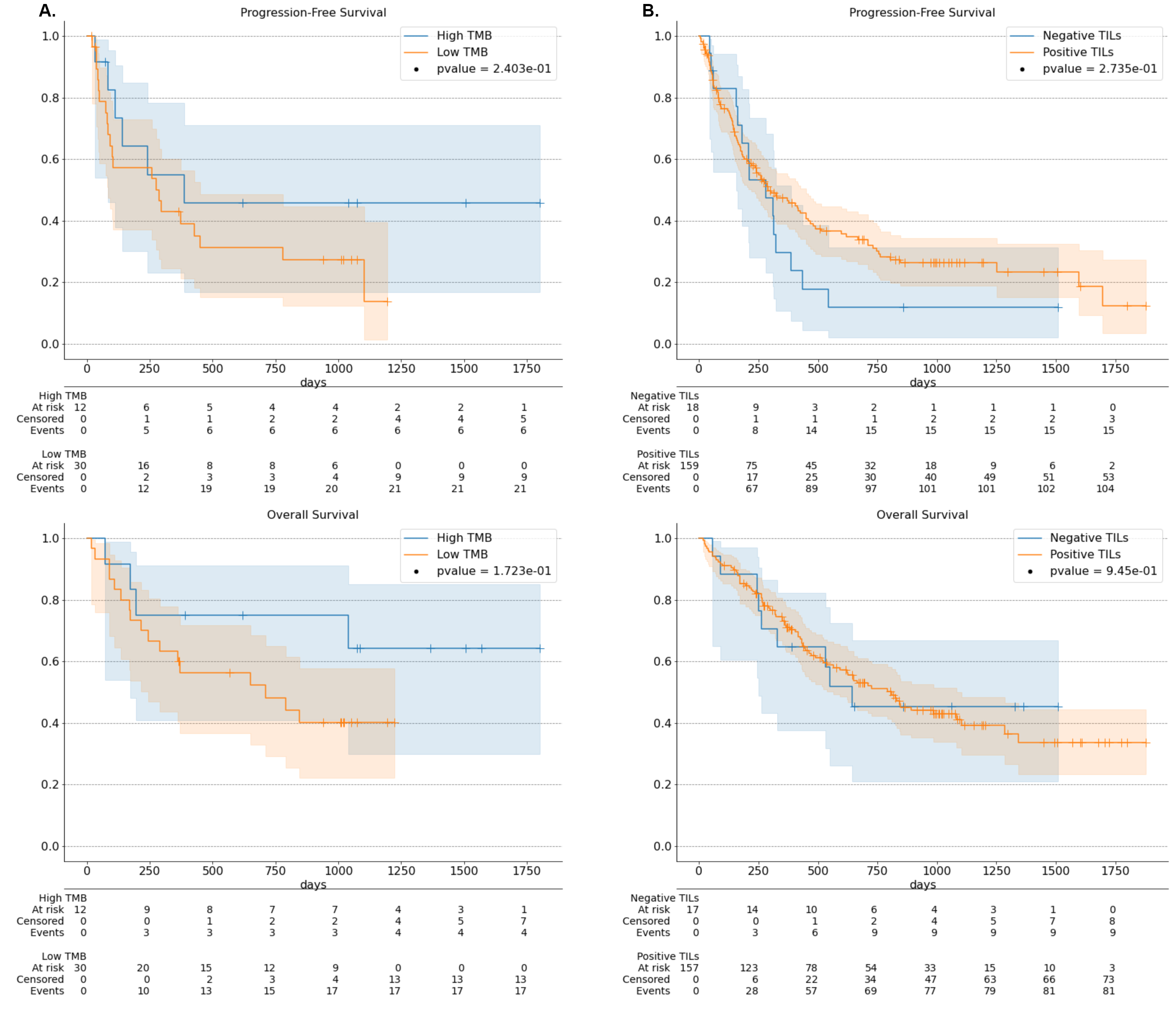

**Figure s3: A.** OS (bottom) and PFS (top) Kaplan-Meier survival curves with 95% confidence interval for the 42 patients with available TMB. Patients are stratified with a threshold of 15 mutations per megabase. Log-rank p-values as well as the number of patients at risk are reported to assess the separation of the survival curves. **B.** OS (bottom) and PFS (top) Kaplan-Meier survival curves with 95% confidence interval for the 174 patients with available TILs status. Patients are stratified with respect to their positive vs negative TILs status. Log-rank p-values as well as the number of patients at risk are reported to assess the separation of the survival curves.

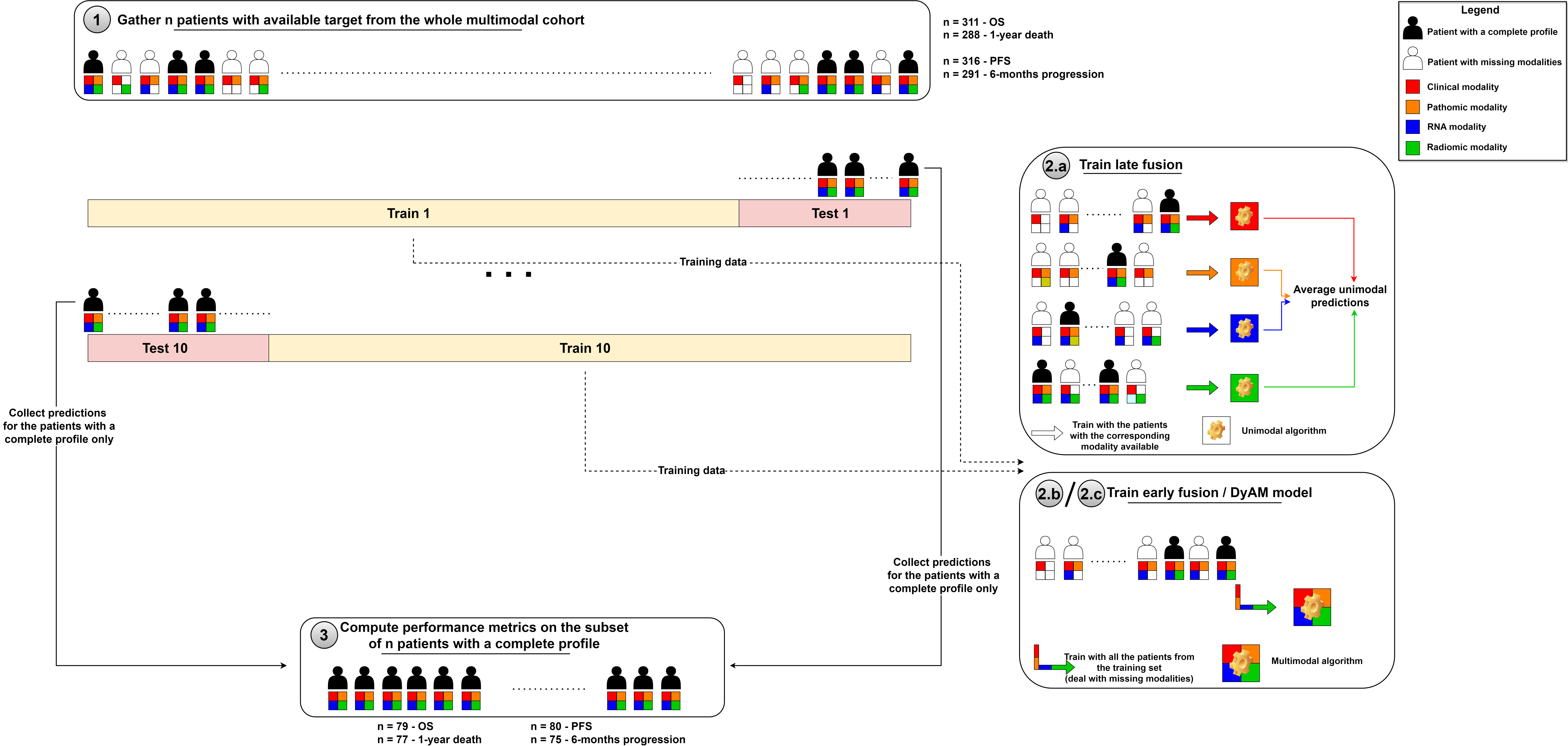

**Figure s4:** Train and test pipeline for all the predictive models. Here the combination with 4 modalities is represented but this scheme is valid for every combination (removing patients from the training set with no modalities available). 1. All the patients with available target (i.e., OS, 1-year death, PFS, or 6-months progression) are collected from the whole multimodal cohort (n=317), they are shuffled and a stratified 10-fold cross-validation scheme is applied. 2. For each fold, data from the training set are used to train the predictive model (pre-processing + learning algorithm), whether it is a unimodal model, a late fusion model, an early fusion model or a DyAM model. The trained predictive model is then applied to the test set of the corresponding fold. 3. Predictions from each test set are collected, considering only those made for the patients with a complete profile. Lastly, performance metrics are computed with this subset of collected predictions.

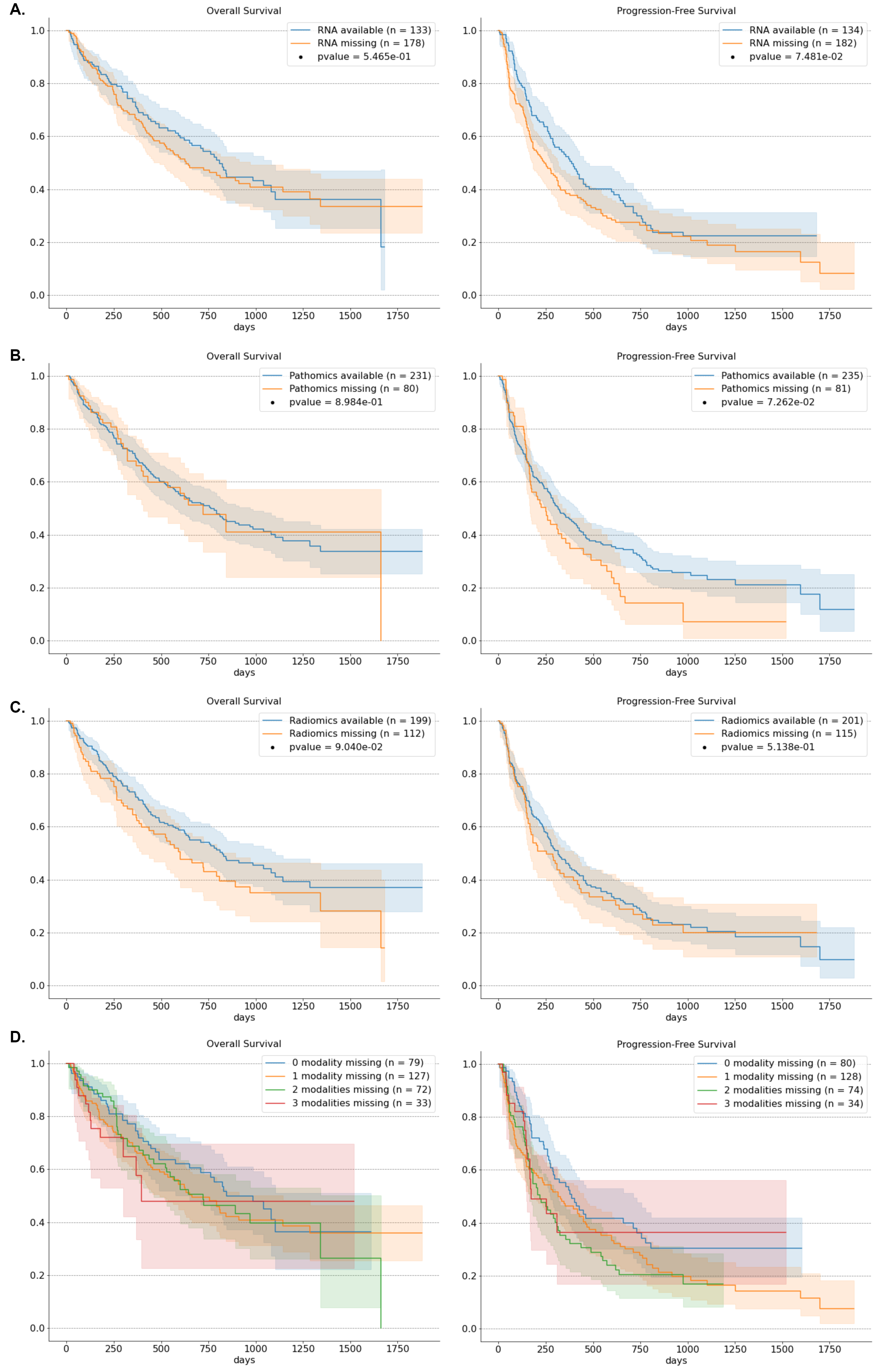

**Figure s5:** OS and PFS Kaplan-Meier survival curves with 95% confidence interval for the whole NSCLC cohort (n=311 for OS and n=316 for PFS). Log-rank p-values are reported to assess the separation of survival curves. **A.** Patients are stratified with respect to the availability of RNA data. **B.** Patients are stratified with respect to the availability of pathomics data. **C.** Patients are stratified with respect to the availability of radiomics data. **D.** Patients are stratified with respect to the number of missing modalities.

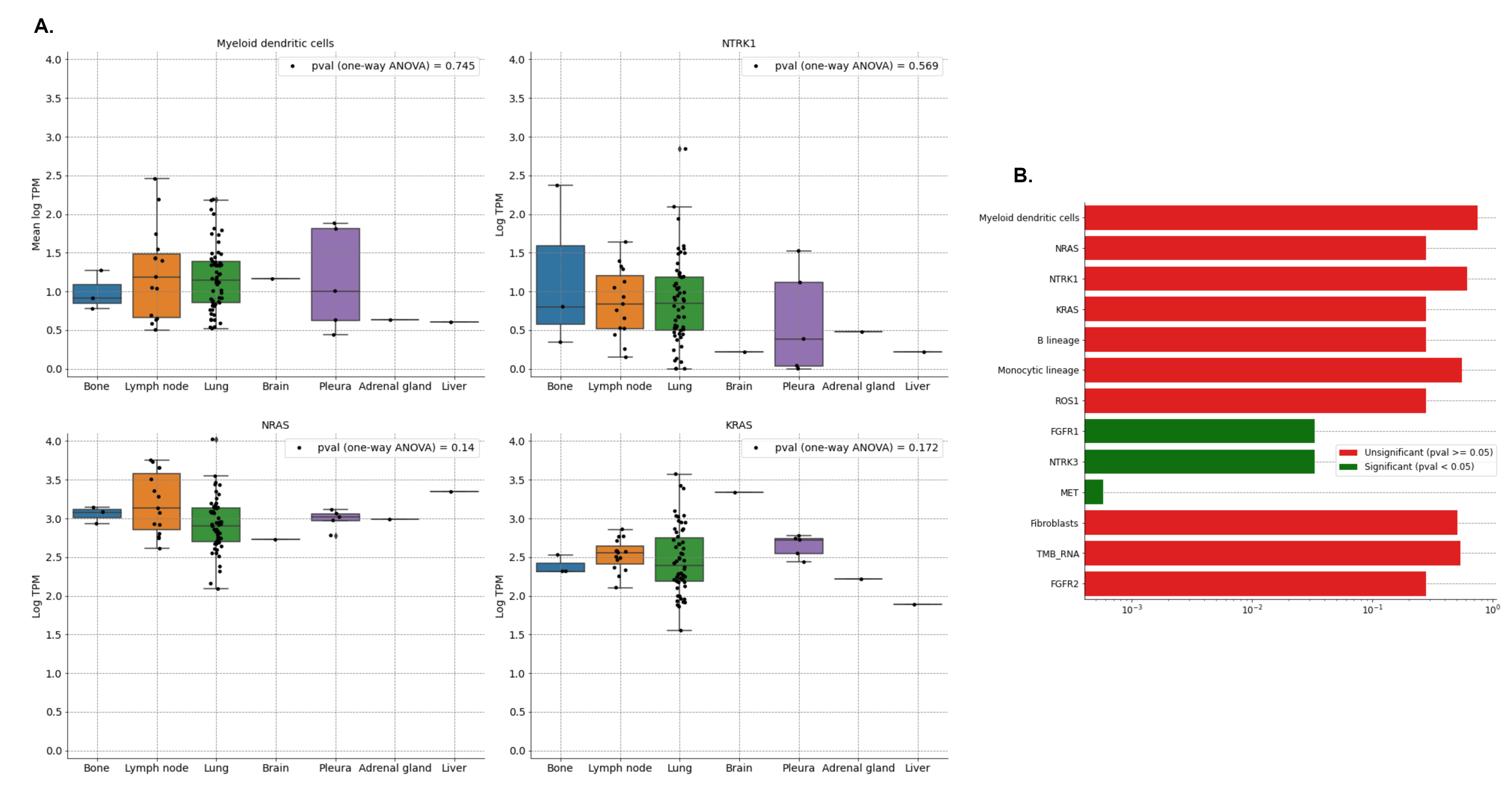

**Figure s6:** Association between the consensus transcriptomic features (i.e., involved in the prediction of overall survival) and the biopsy site (n=84 patients). One-way ANOVA tests are used to test for statistically significant differences between the means of the transcriptomic feature in the different biopsy sites. **A.** Distribution of the four most important transcriptomic features in the different biopsy sites. **B.** One-way ANOVA p-values for each of the 13 consensus transcriptomic features, displayed with a bar plot in log-scale. P-values were corrected with Benjamini-Hochberg procedure (FDR controlled at level $\alpha=0.05$).

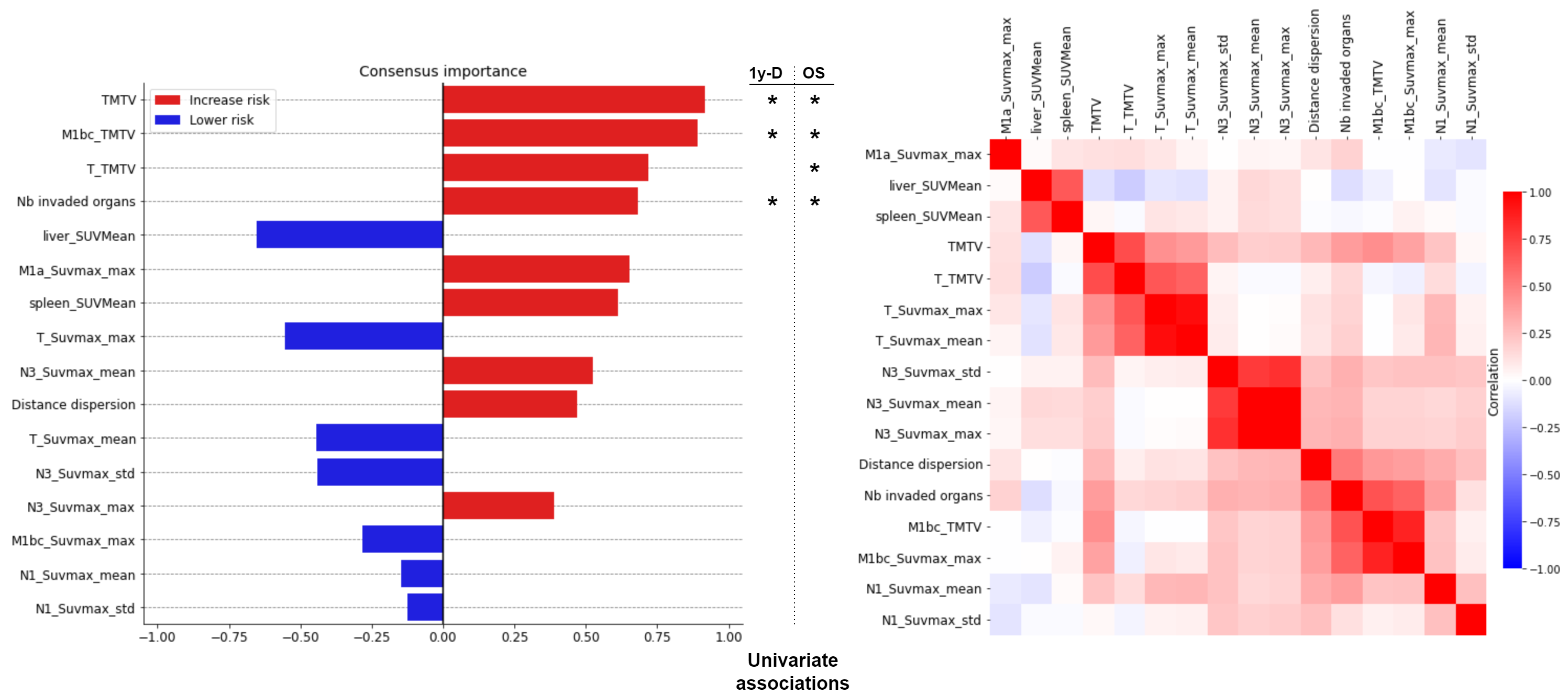

**Figure s7:** Feature importance ranking for the prediction of overall survival (OS and 1-year death) with radiomic modality (left) and heatmap of Spearman correlations between consensus radiomic features (right). Logistic regression was not taken into account for computing this ranking since its AUC was lower than 0.5. Features that are significantly associated with 1-year death (permutation test with univariate AUCs) after Benjamini-Hochberg (BH) correction ($\alpha=0.05$) are annotated with a * on the left side, while features that are significantly associated with OS (permutation test with univariate C-index) after BH correction are annotated with a * on the right side.

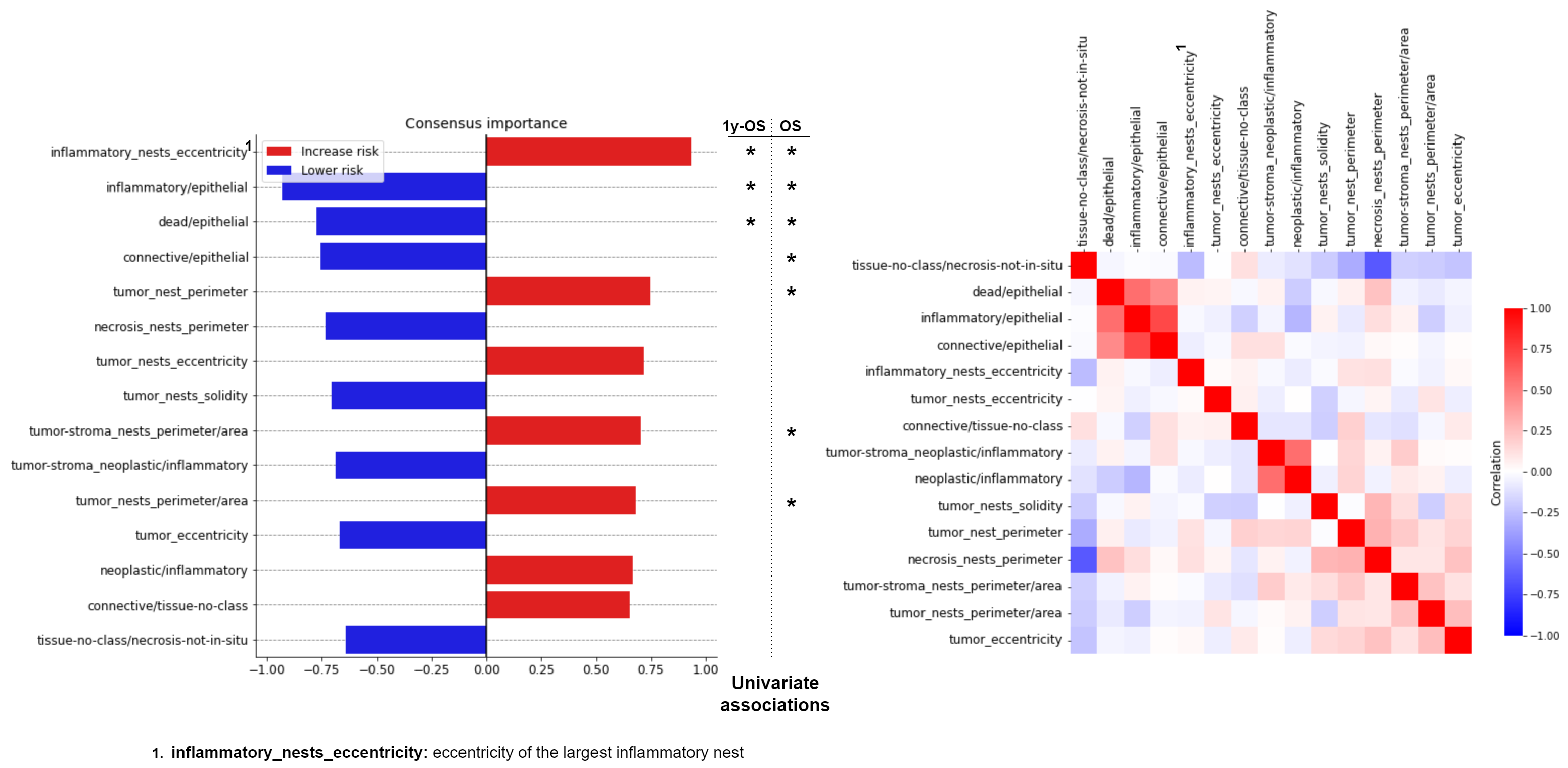

**Figure s8:** Feature importance ranking for the prediction of overall survival (OS and 1-year death) with pathomic modality (left) and heatmap of Spearman correlations between consensus pathomic features (right). Features that are significantly associated with 1-year death (permutation test with univariate AUCs) after Benjamini-Hochberg (BH) correction ($\alpha=0.05$) are annotated with a * on the left side, while features that are significantly associated with OS (permutation test with univariate C-index) after BH correction are annotated with a * on the right side.

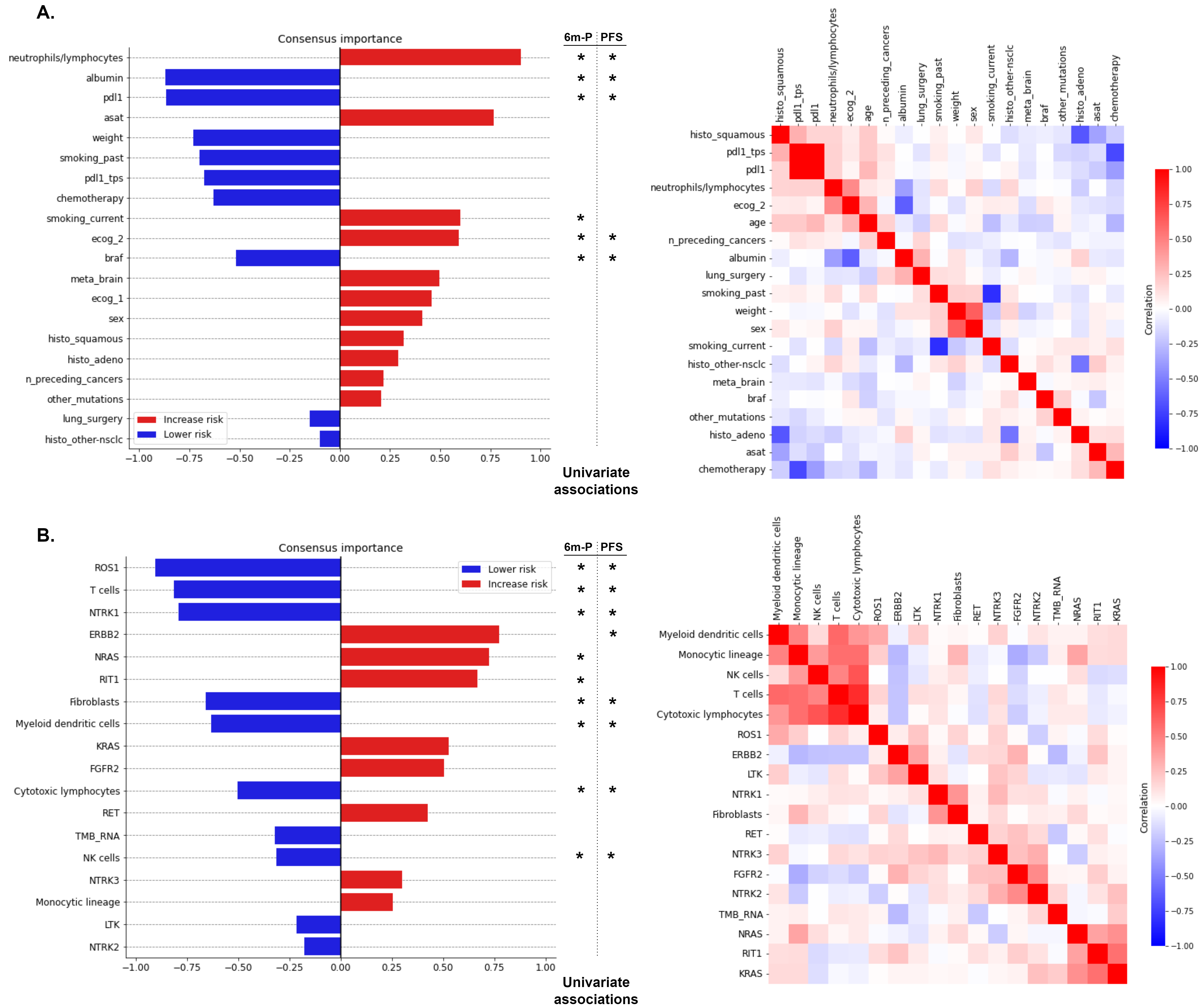

**Figure s9:** Feature importance ranking for the prediction of progression-free survival, obtained by aggregating the SHAP values collected from both tasks (PFS and 6-months progression) and both approaches (linear and tree ensemble methods) (supplementary methods 2.c). Features that are significantly associated with 6-months progression (permutation test with univariate AUCs) after Benjamini-Hochberg (BH) correction ($\alpha=0.05$) are shown with a * on the left side, while features that are significantly associated with PFS (permutation test with univariate C-index) after BH correction are annotated with a * on the right side. **A.** Consensus feature importance ranking for the clinical data modality (left) and heatmap of correlations between consensus clinical features (right). Correlations were evaluated by Spearman correlation coefficients (for continuous feature vs continuous feature), AUCs rescaled to $[-1, 1]$(for continuous feature vs binary categorical feature), or Matthews correlation coefficient (for binary categorical feature vs binary categorical feature). **B.** Consensus feature importance ranking for the RNA data modality (left) and heatmap of Spearman correlations between consensus RNA features (right).

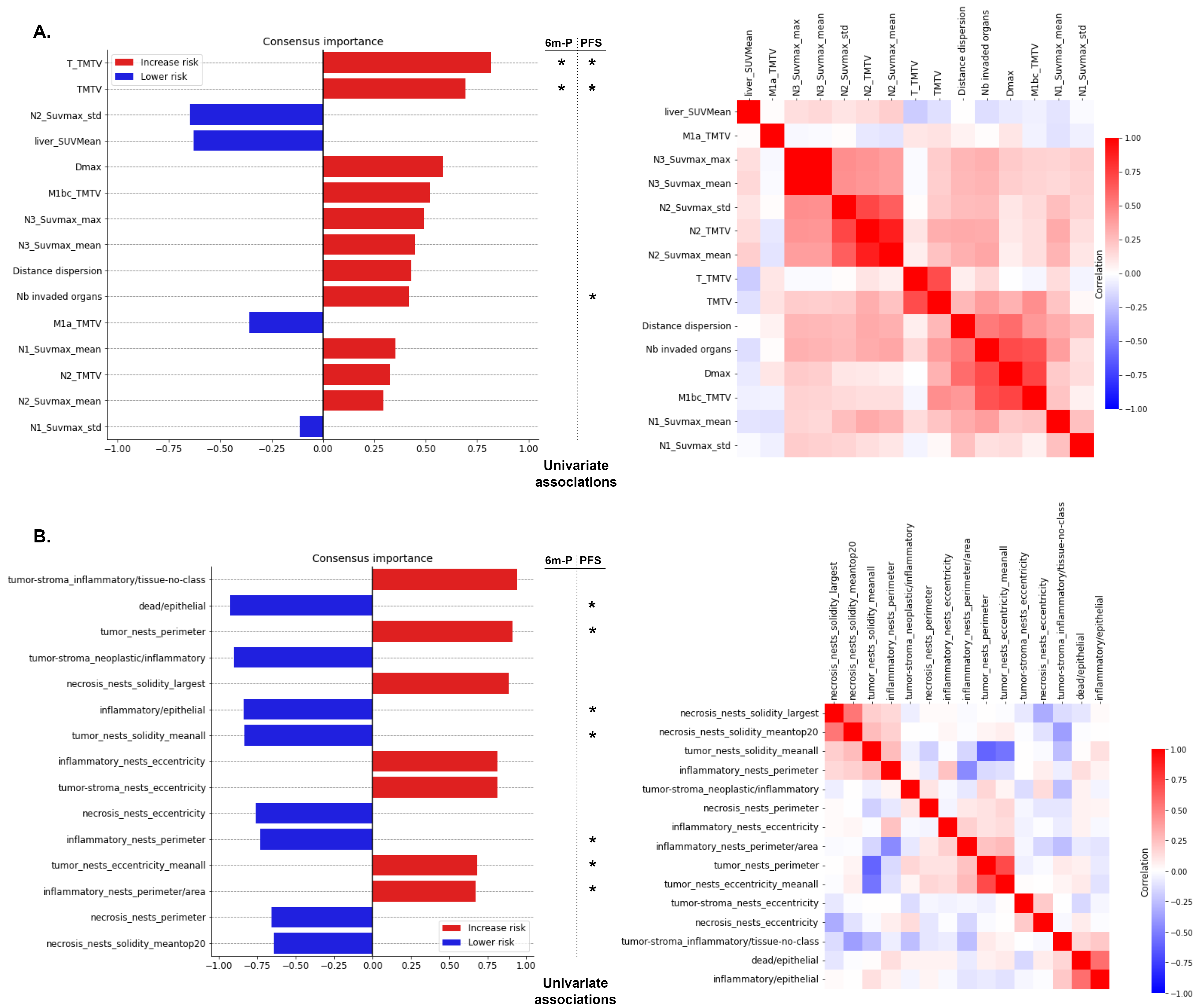

**Figure s10:** Feature importance ranking for the prediction of progression-free survival, obtained by aggregating the SHAP values collected from both tasks (PFS and 6-months progression) and both approaches (linear and tree ensemble methods) (supplementary methods 2.c). **A.** Consensus feature importance ranking for the radiomic data modality (left) and heatmap of Spearman correlations between consensus radiomic features (right). **B.** Consensus feature importance ranking for the pathomic data modality (left) and heatmap of Spearman correlations between consensus pathomic features (right).

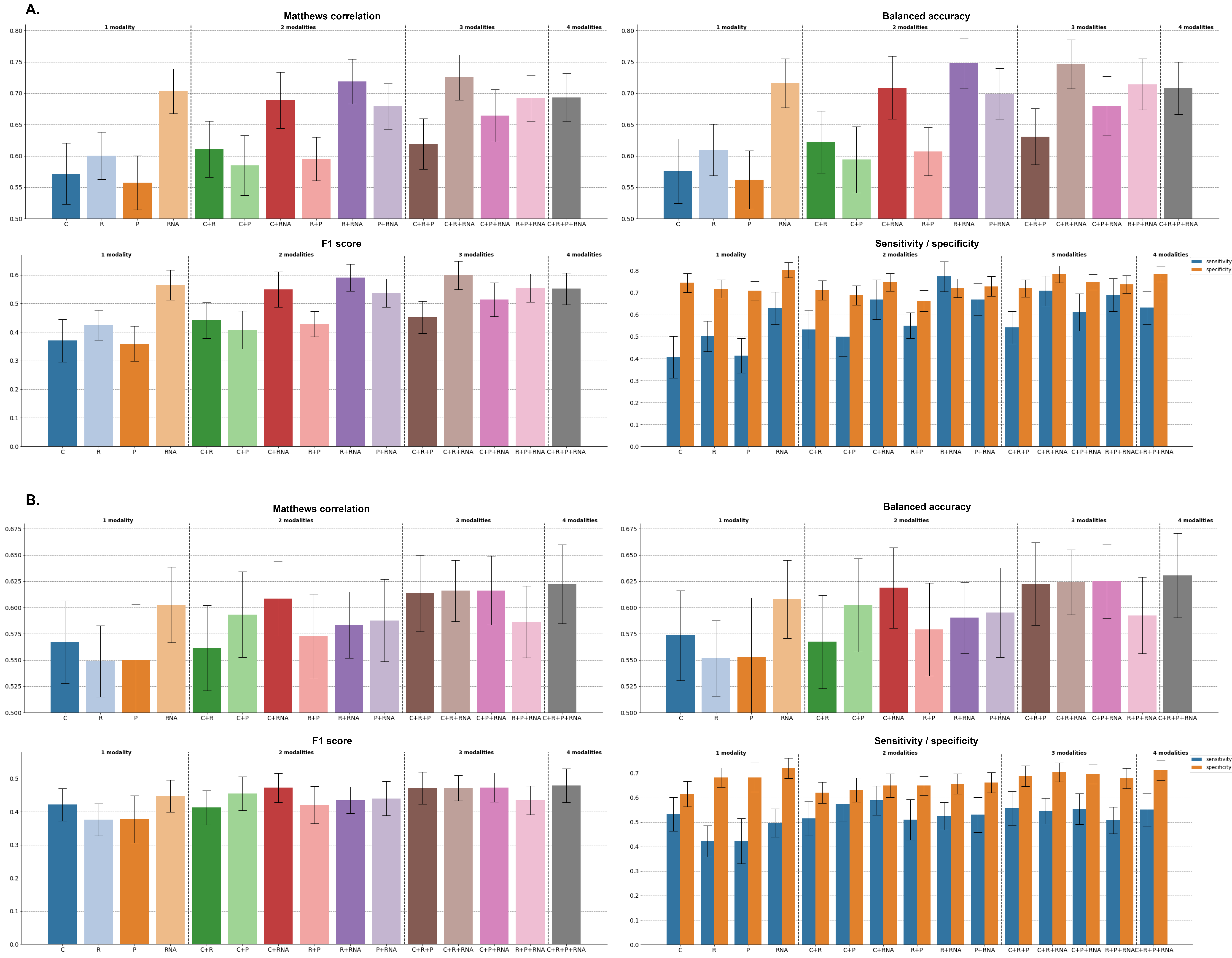

**Figure s11:** Binary classification performance of all the possible multimodal combinations, with a late fusion strategy and XGBoost for 1-year death and 6-months progression prediction (C: clinical, R: radiomic, P: pathomic, RNA). The bar height corresponds to the performance metric (balanced Accuracy, Matthews correlation coefficient, F1 score, and sensitivity and specificity) averaged across the 100 cross-validation schemes, and the error bar corresponds to ± 1 standard deviation, estimated across the 100 cross-validation schemes. **A.** Binary metrics associated with the prediction of 1-year death with XGBoost algorithms and estimated with n=77 patients**. B.** Binary metrics associated with the prediction of 6-months progression with XGBoost algorithms and estimated with n=75 patients.

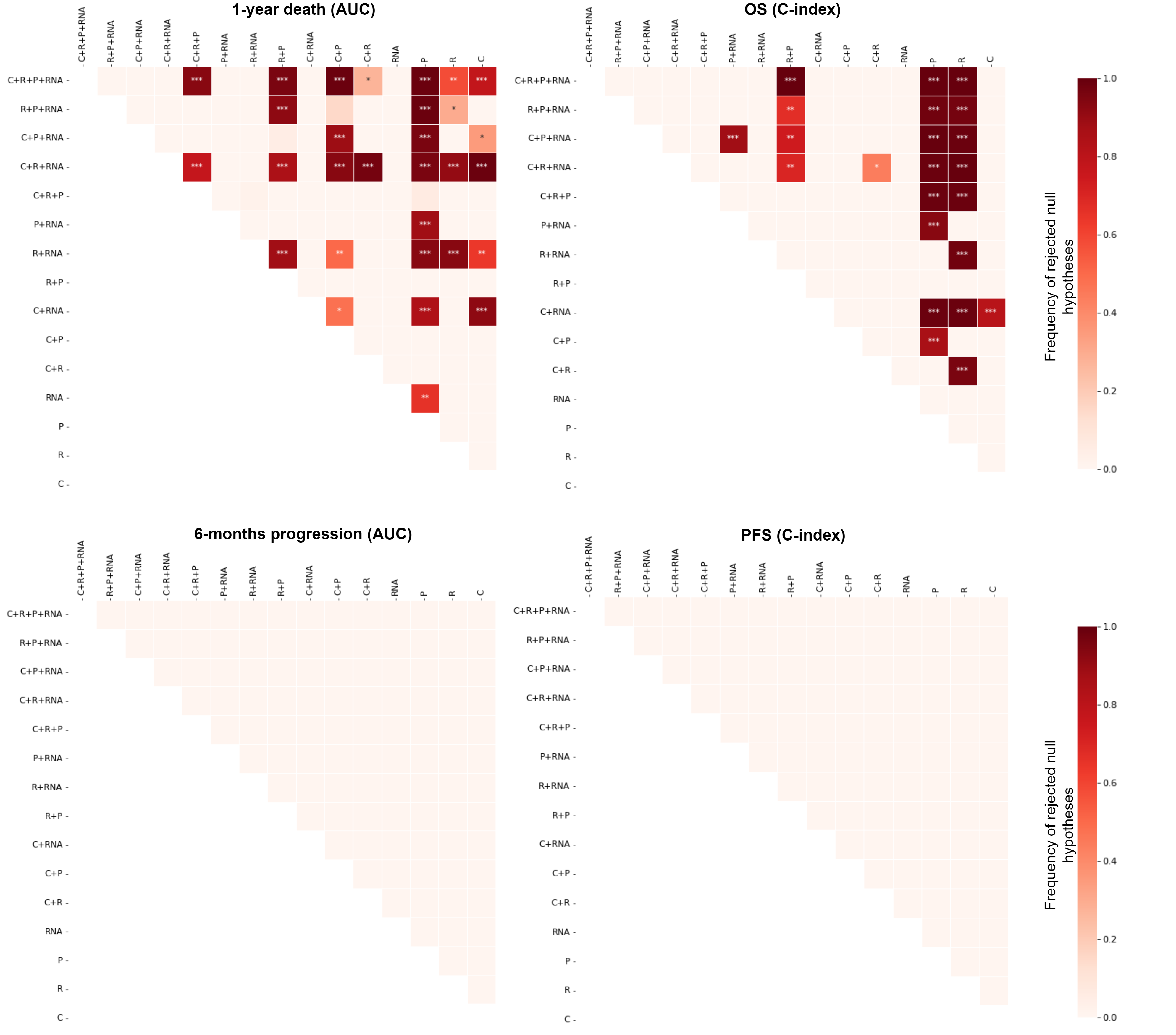

**Figure s12:** Paired permutation tests (21) to test for statistically significant differences between the performances (AUC or C-index) of the different multimodal combinations for the late fusion strategy with tree ensemble methods (supplementary methods section 4.a). Each element of the heatmap corresponds to the test for the superiority of the row multimodal combination over the column multimodal combination (e.g., the top right corner of the heatmap corresponds to the test of the superiority of the combination with the four modalities over the clinical model). The lower parts of the heatmaps are not shown since they contain no significant comparison. The color scale corresponds to the frequency of statistically significant tests across the 100 cross-validation schemes after Benjamini-Hochberg correction (FDR controlled at level $\alpha=0.05).$ * corresponds to 25 to 50% of significant tests, ** corresponds to 50 to 75% of significant tests, and *** corresponds to more than 75% of significant tests.

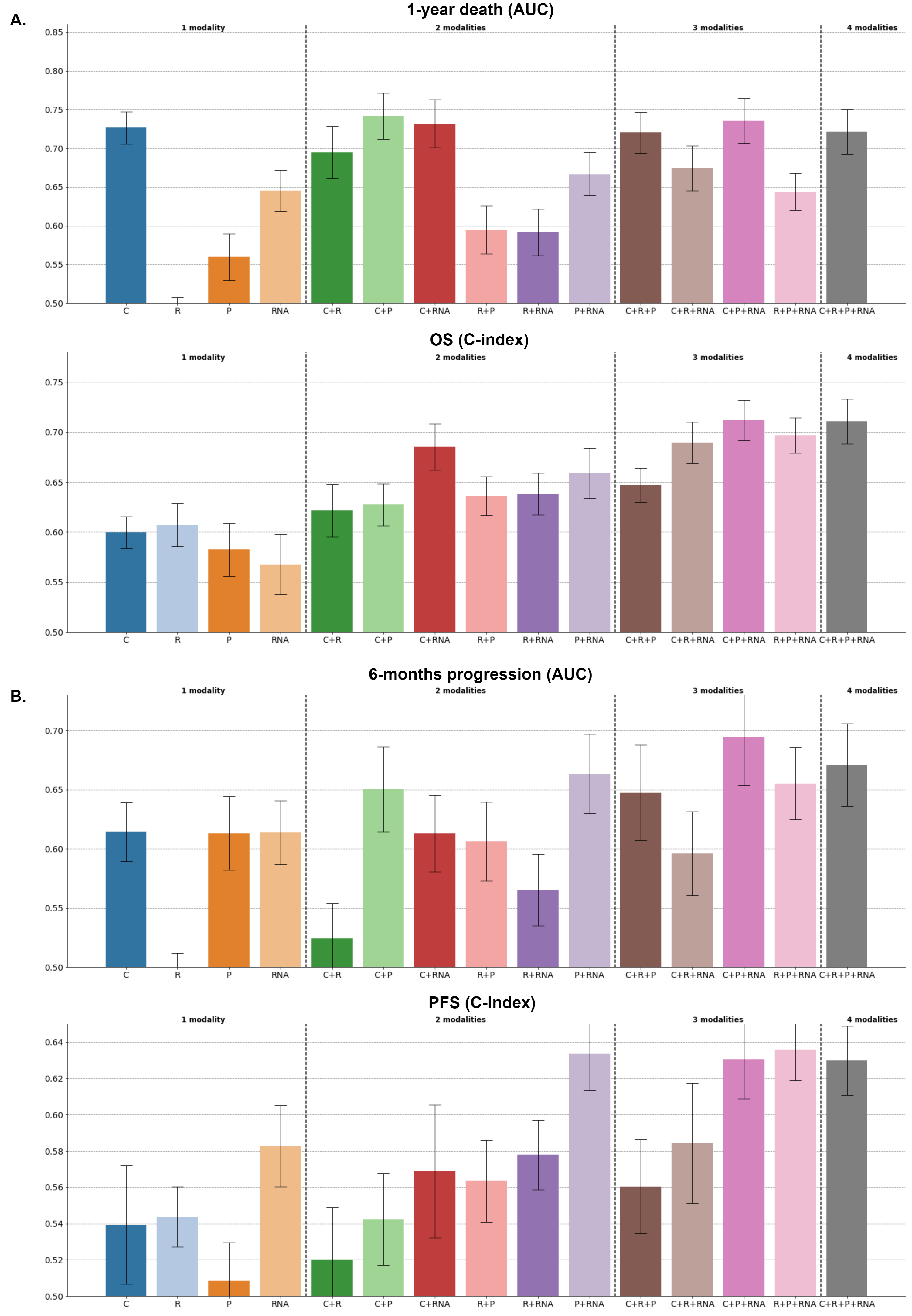

**Figure s13:** Performance of all the possible multimodal combinations, with a late fusion strategy and linear methods (C: clinical, R: radiomic, P: pathomic, RNA). **A.** ROC AUCs associated with the prediction of 1-year death with penalized logistic regression algorithms (top) and estimated with n=77 patients. C-indexes associated with the prediction of OS with penalized Cox’s regression algorithms (bottom) and estimated with n=79 patients. **B**. ROC AUCs associated with the prediction of 6-months progression with penalized logistic regression algorithms (top) and estimated with n=75 patients. C-indexes associated with the prediction of PFS with penalized Cox’s regression (bottom) and estimated with n=80 patients.

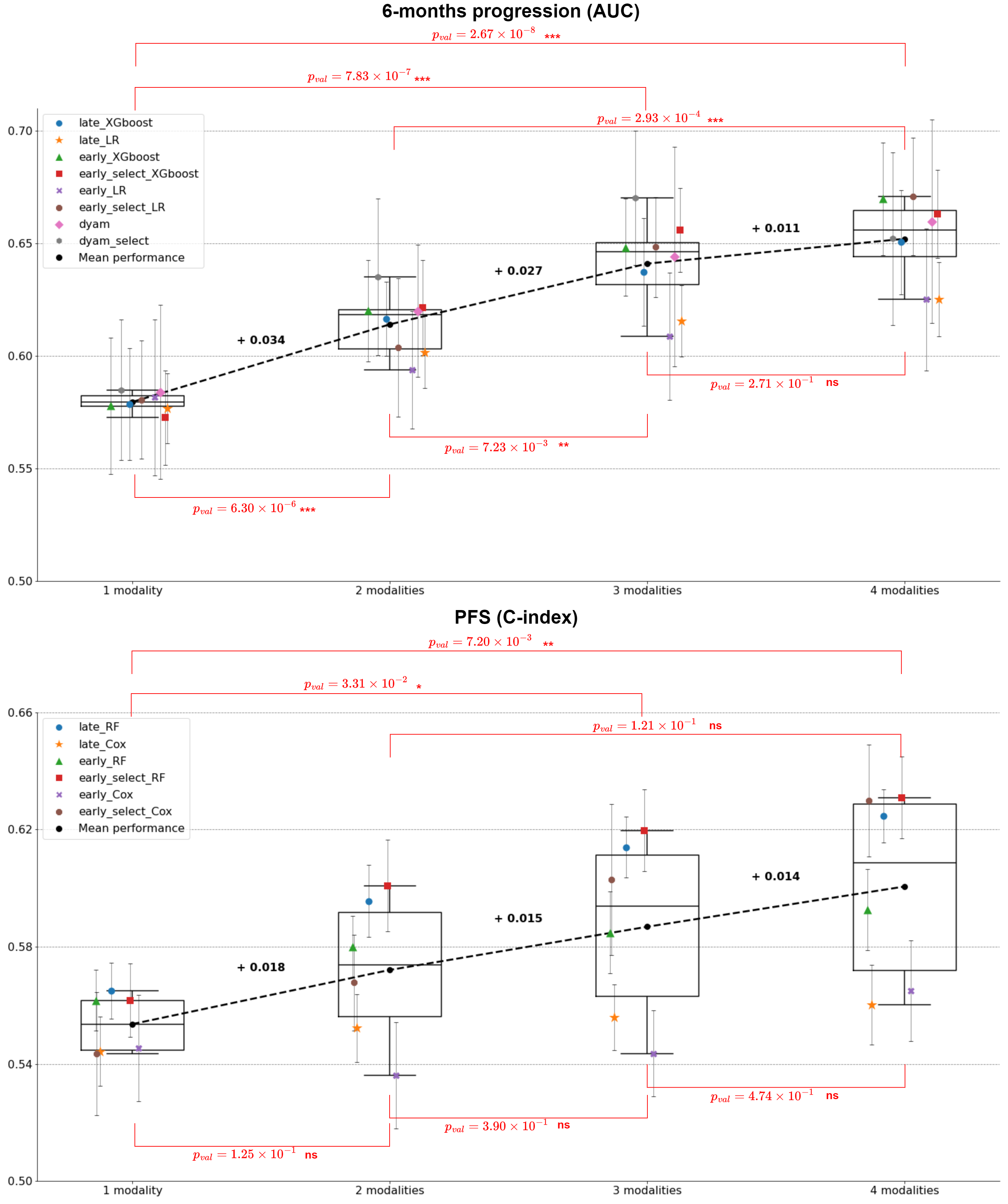

**Figure s14:** Average performance across all models with 1 modality, 2 modalities, 3 modalities and 4 modalities for 6-months progression (top) and PFS (bottom). Markers and error bars correspond to the mean average performance and ± 1 standard deviation respectively, estimated across the 100 cross-validation schemes. Mean increases are represented with dashed lines and bold annotations. Red annotations correspond to Student’s t test p-values to compare the different numbers of integrated modalities (e.g., 1 modality vs 2 modalities).

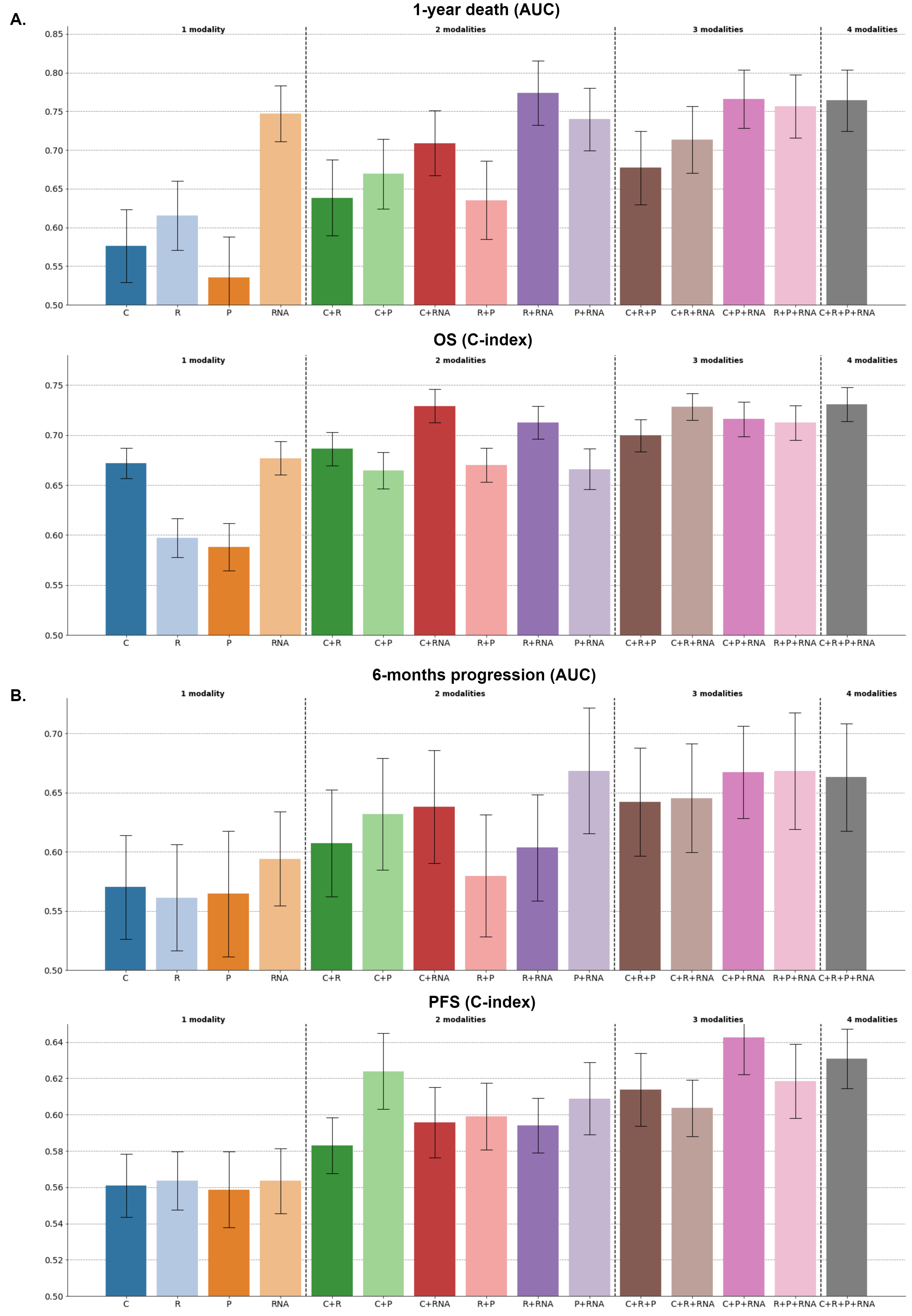

**Figure s15:** Performance of all the possible multimodal combinations with an early fusion strategy, tree ensemble methods, and a preliminary feature selection step (C: clinical, R: radiomic, P: pathomic, RNA). **A.** ROC AUCs associated with the prediction of 1-year death with XGBoost algorithms (top) and estimated with n=77 patients. C-indexes associated with the prediction of OS with Random Forest Survival algorithms (bottom) and estimated with n=79 patients. **B**. ROC AUCs associated with the prediction of 6-months progression with XGBoost algorithms (top) and estimated with n=75 patients. C-indexes associated with the prediction of PFS with Random Survival Forest algorithms (bottom) and estimated with n=80 patients.

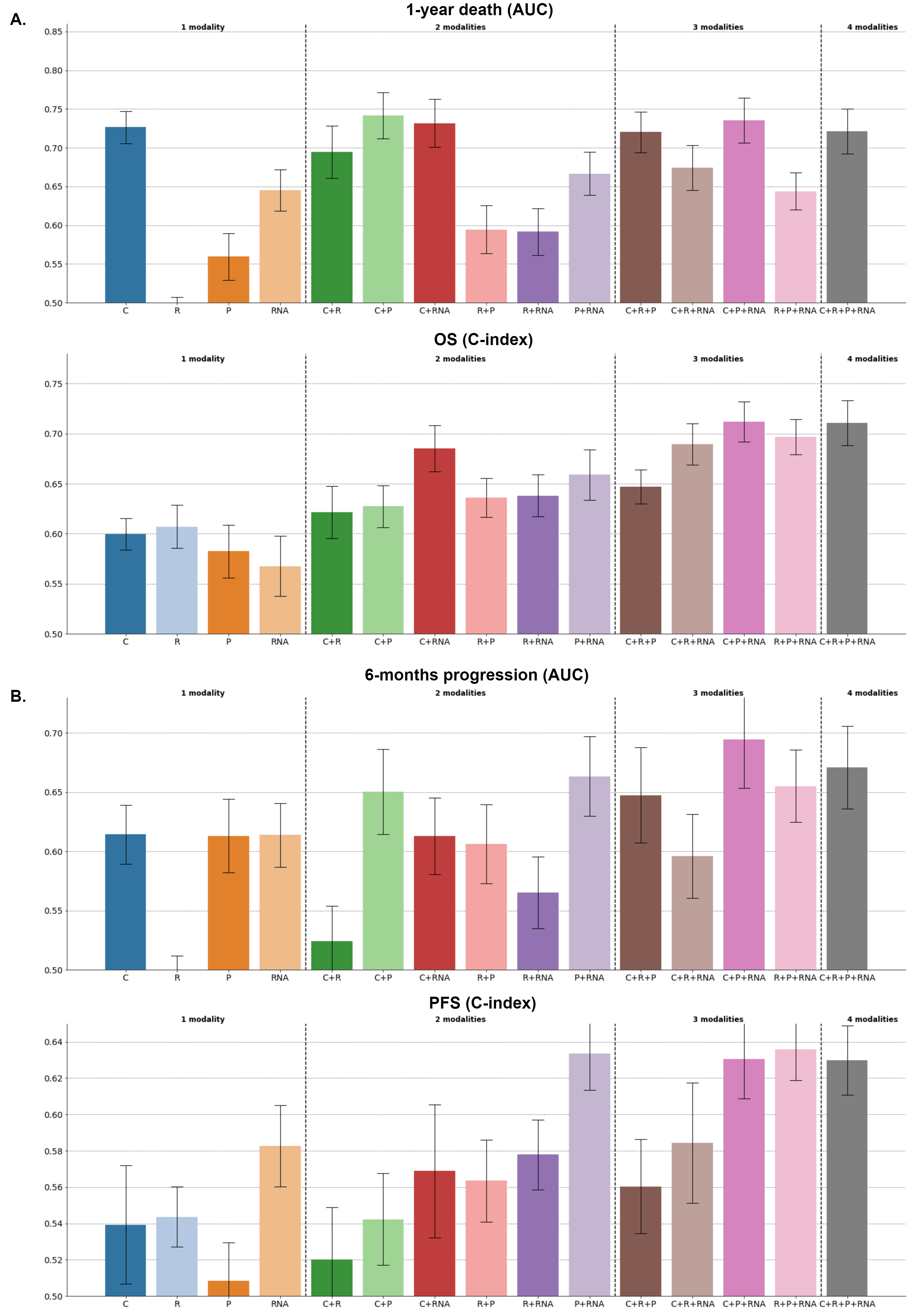

**Figure s16:** Performance of all the possible multimodal combinations with an early fusion strategy, linear methods, and a preliminary feature selection step (C: clinical, R: radiomic, P: pathomic, RNA). **A.** ROC AUCs associated with the prediction of 1-year death with penalized logistic regression algorithms (top) and estimated with n=77 patients. C-indexes associated with the prediction of OS with penalized Cox’s regression algorithms (bottom) and estimated with n=79 patients. **B**. ROC AUCs associated with the prediction of 6-months progression with penalized logistic regression algorithms (top) and estimated with n=75 patients. C-indexes associated with the prediction of PFS with penalized Cox’s regression (bottom) and estimated with n=80 patients.

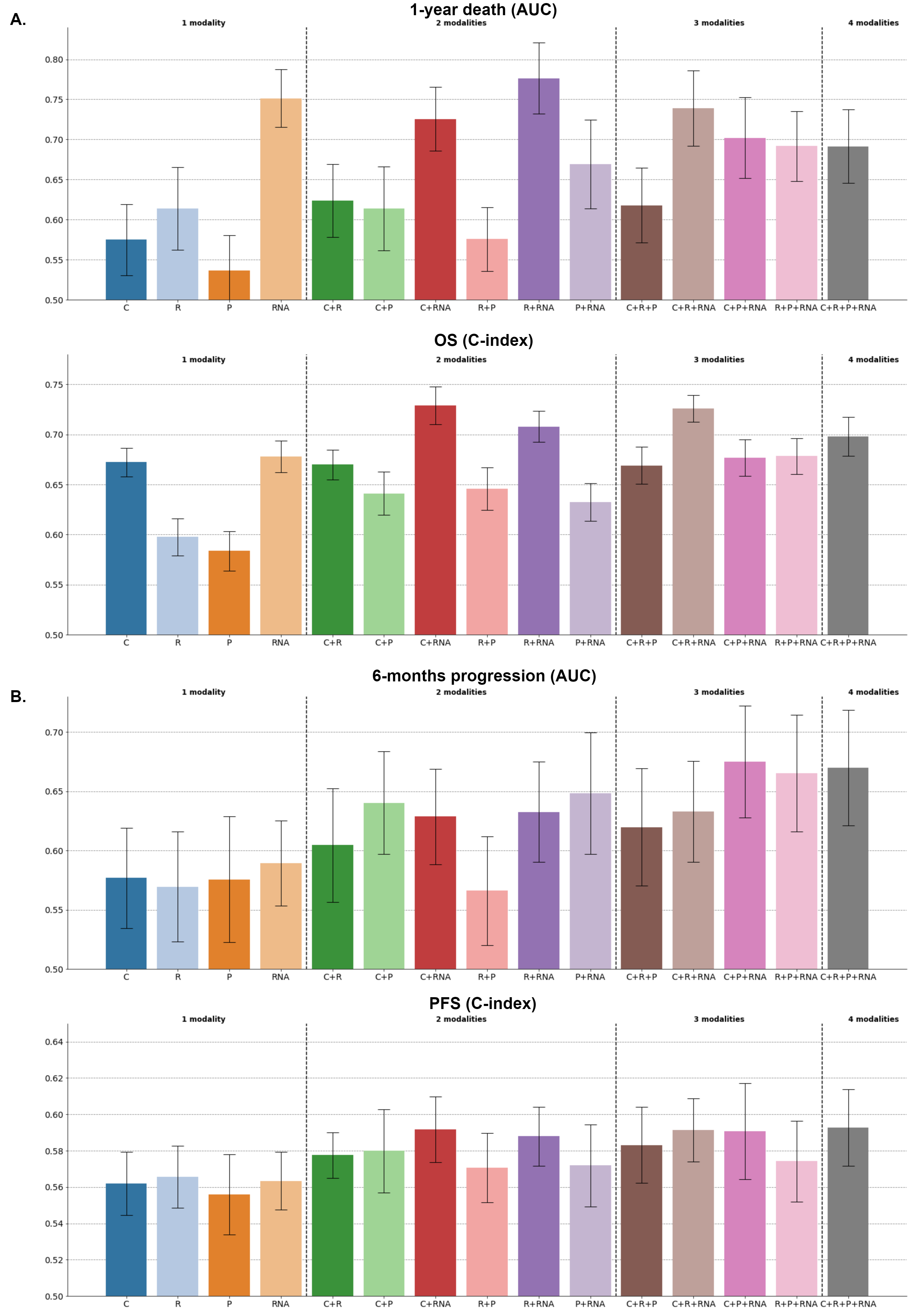

**Figure s17:** Performance of all the possible multimodal combinations with an early fusion strategy and tree ensemble methods (C: clinical, R: radiomic, P: pathomic, RNA). **A.** ROC AUCs associated with the prediction of 1-year death with XGBoost algorithms (top) and estimated with n=77 patients. C-indexes associated with the prediction of OS with Random Forest Survival algorithms (bottom) and estimated with n=79 patients. **B**. ROC AUCs associated with the prediction of 6-months progression with XGBoost algorithms (top) and estimated with n=75 patients. C-indexes associated with the prediction of PFS with Random Survival Forest algorithms (bottom) and estimated with n=80 patients.

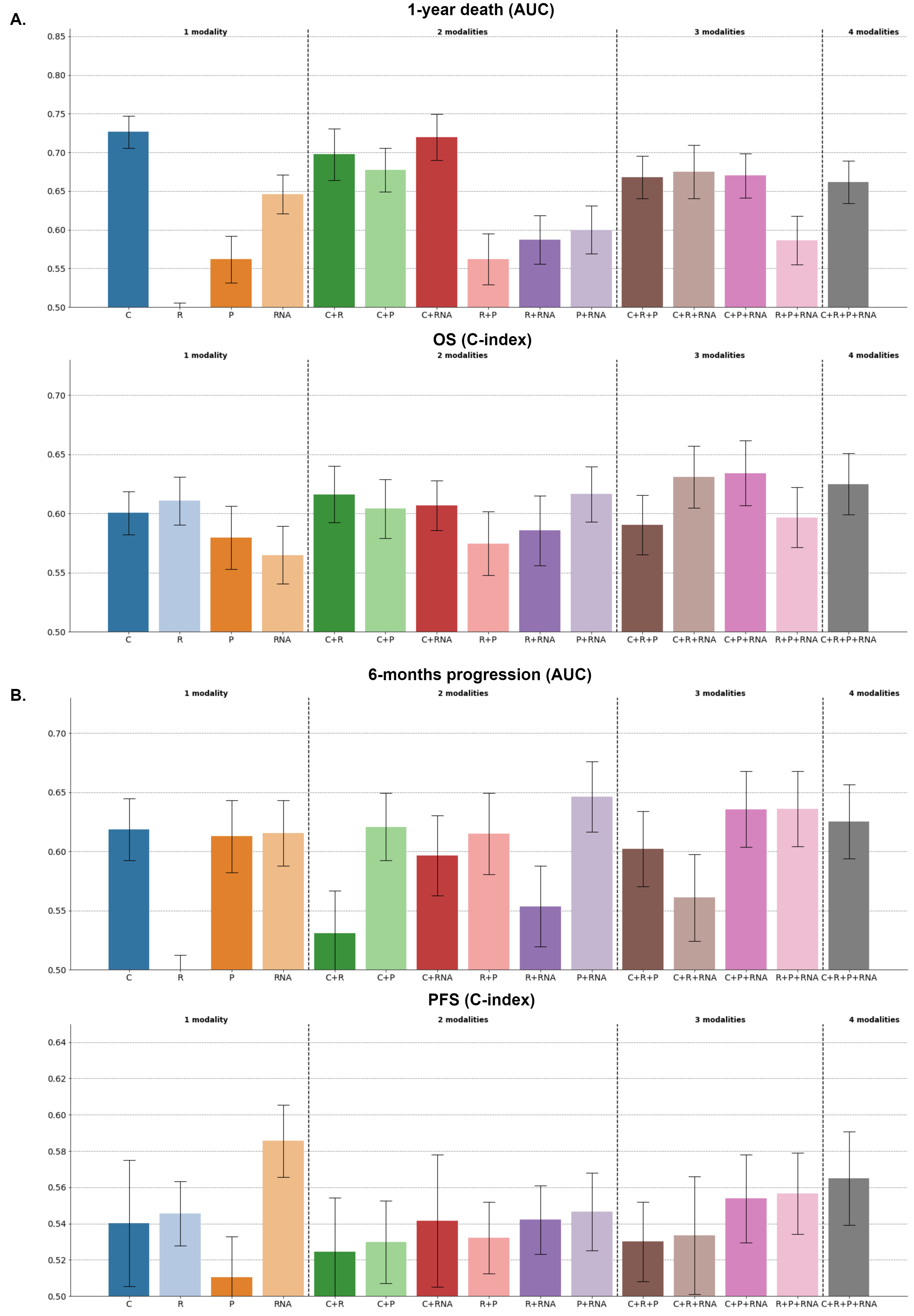

**Figure s18:** Performance of all the possible multimodal combinations with an early fusion strategy and linear methods (C: clinical, R: radiomic, P: pathomic, RNA). **A.** ROC AUCs associated with the prediction of 1-year death with penalized logistic regression algorithms (top) and estimated with n=77 patients. C-indexes associated with the prediction of OS with penalized Cox’s regression algorithms (bottom) and estimated with n=79 patients. **B**. ROC AUCs associated with the prediction of 6-months progression with penalized logistic regression algorithms (top) and estimated with n=75 patients. C-indexes associated with the prediction of PFS with penalized Cox’s regression (bottom) and estimated with n=80 patients.

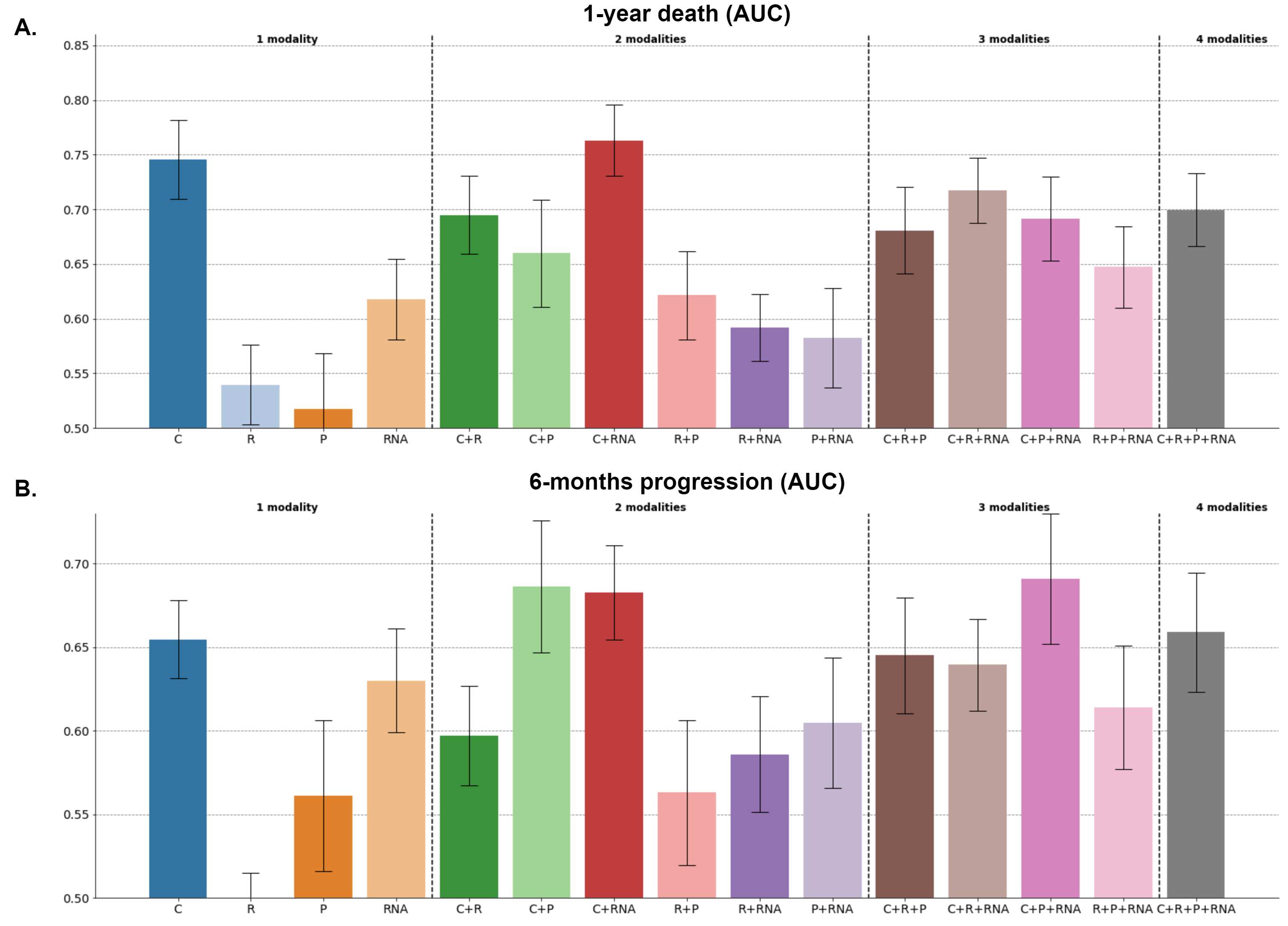

**Figure s19:** Performance of all the possible multimodal combinations with DyAM strategy (C: clinical, R: radiomic, P: pathomic, RNA). **A.** ROC AUCs associated with the prediction of 1-year death and estimated with n=77 patients. **B.** ROC AUCs associated with the prediction of 6-months progression and estimated with n=75 patients.

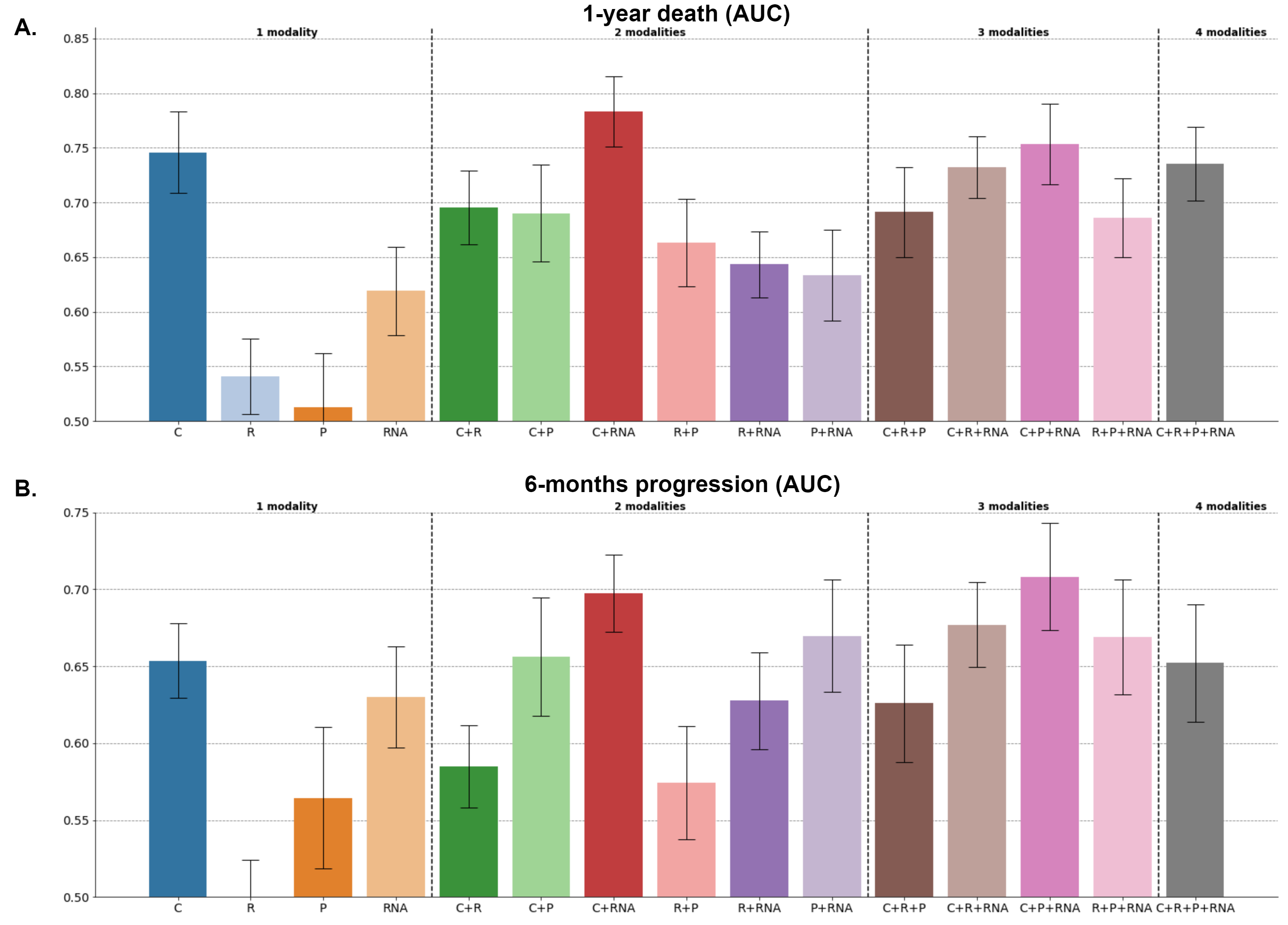

**Figure s20:** Performance of all the possible multimodal combinations with DyAM strategy and a preliminary feature selection step. **A.** ROC AUCs associated with the prediction of 1-year death and estimated with n=77 patients. **B.** ROC AUCs associated with the prediction of 6-months progression and estimated with n=75 patients.

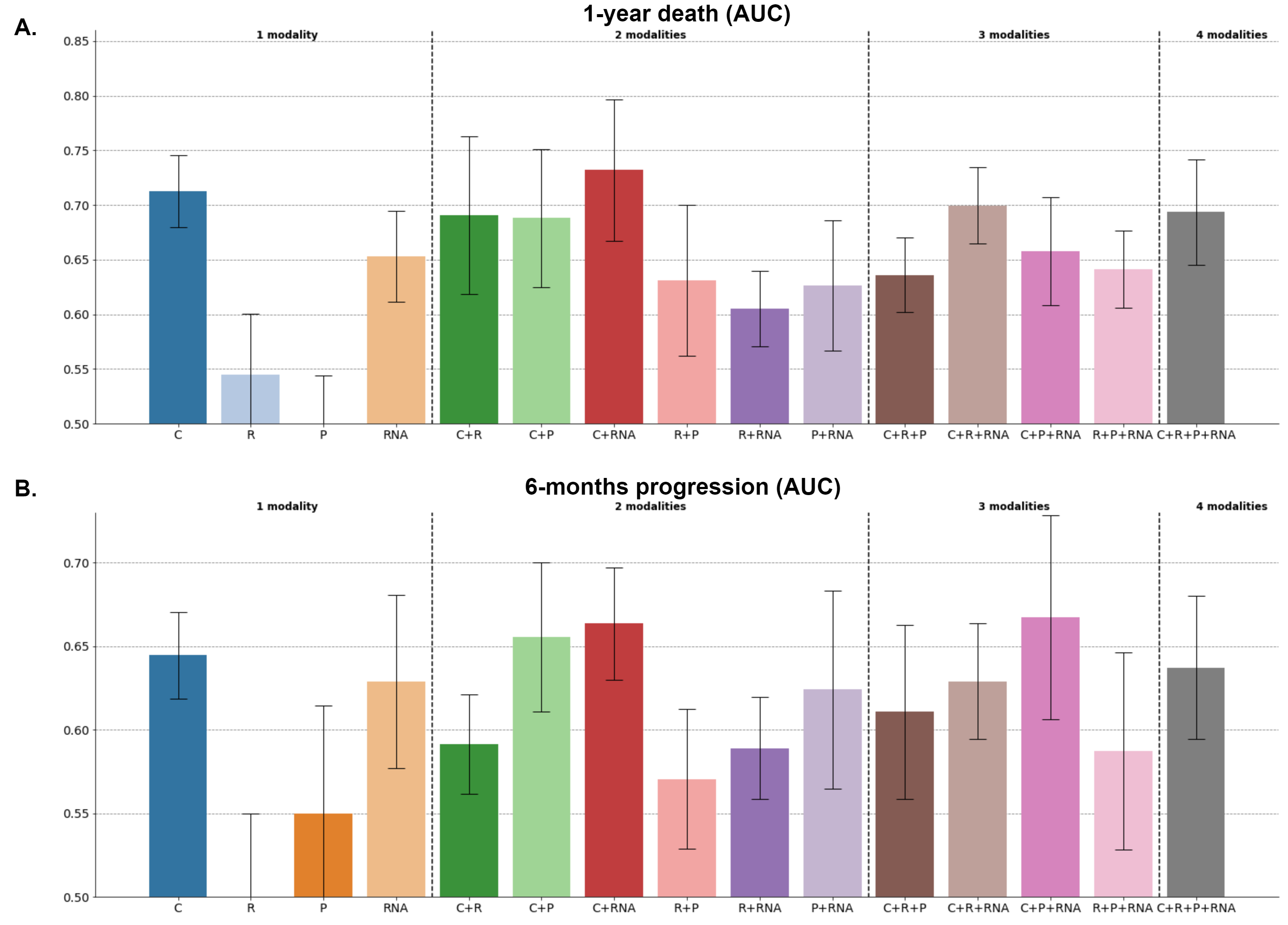

**Figure s21:** Performance of all the possible multimodal combinations with DyAM strategy. The learning rate and the L2 regularization strength were tuned with a grid-search strategy and a nested-cross validation scheme. The experiment was repeated 10 times instead of 100, due to computational constraints. **A.** ROC AUCs associated with the prediction of 1-year death and estimated with n=77 patients. **B.** ROC AUCs associated with the prediction of 6-months progression and estimated with n=75 patients.

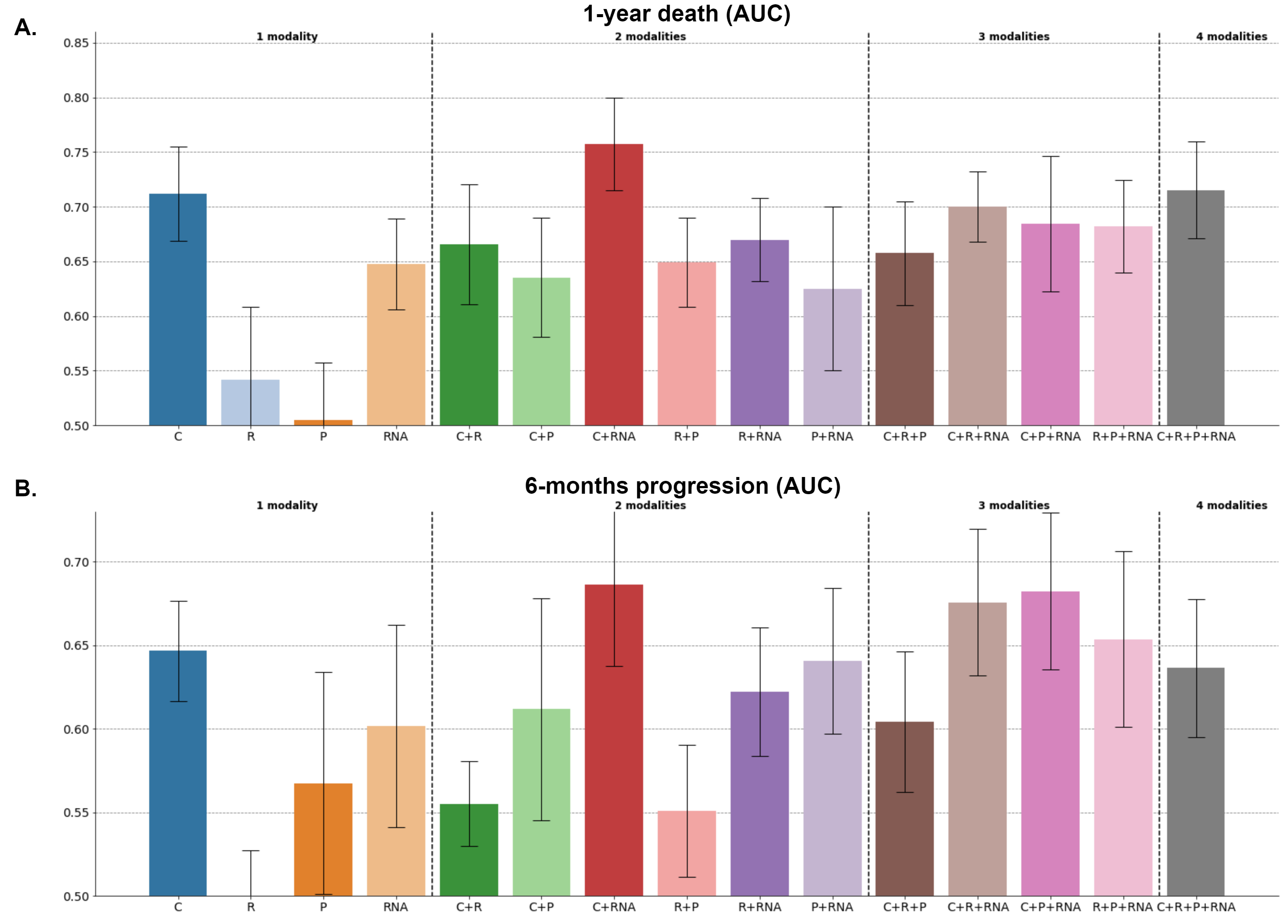

**Figure s22:** Performance of all the possible multimodal combinations with DyAM strategy and a preliminary feature selection step. The learning rate and the L2 regularization strength were tuned with a grid-search strategy and a nested-cross validation scheme. The experiment was repeated 10 times instead of 100, due to computational constraints. **A.** ROC AUCs associated with the prediction of 1-year death and estimated with n=77 patients. **B.** ROC AUCs associated with the prediction of 6-months progression and estimated with n=75 patients.

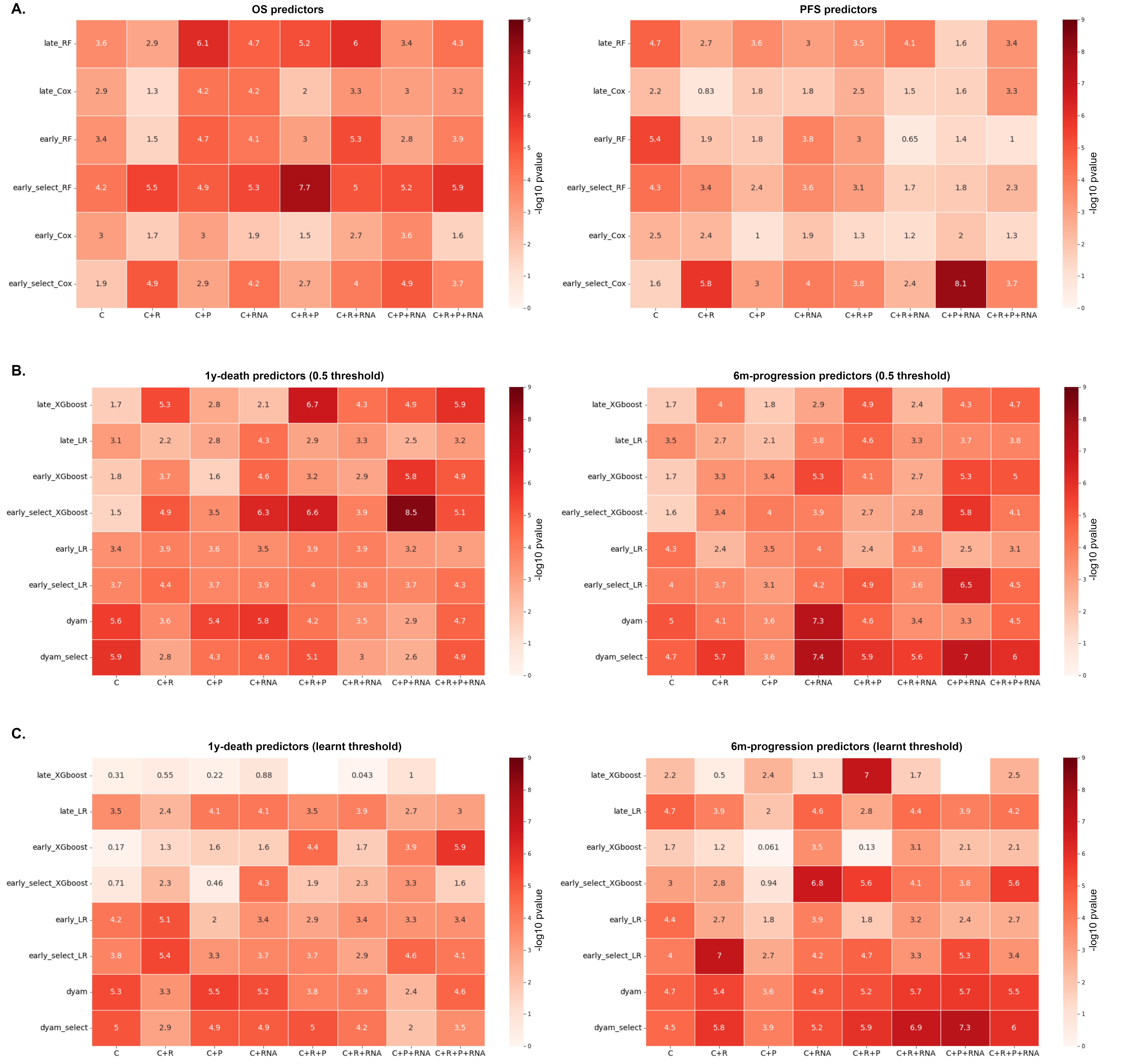

**Figure s23:** Log-rank p-values ($-\log_{10} (p_{val}))$ for the stratification of patients into high-risk and low-risk groups for overall survival, for different predictive tasks (n=265 patients with the 4 targets available for a fair comparison). Only the combinations that include the clinical modality are compared. **A.** Log-rank p-values for models trained to predict OS (left) and models trained to predict PFS (right). Risk-group assignments were determined based on thresholds learned with the model’s predictions collected on the training sets of the cross-validation schemes. For PFS models, thresholds were optimized to stratify PFS but were then applied to stratify OS (see supplementary methods). **B.** Log-rank p-values for models trained to predict 1-year death (left) and models trained to predict 6-months progression (right). Risk-group assignments were determined based on a 0.5 threshold. **C.** Log-rank p-values for models trained to predict 1-year death (left) and models trained to predict 6-months progression (right). Risk-group assignments were determined based on thresholds learned with the model’s predictions collected on the training sets of the cross-validation schemes. For PFS models, thresholds were optimized to stratify PFS but were then applied to stratify OS.

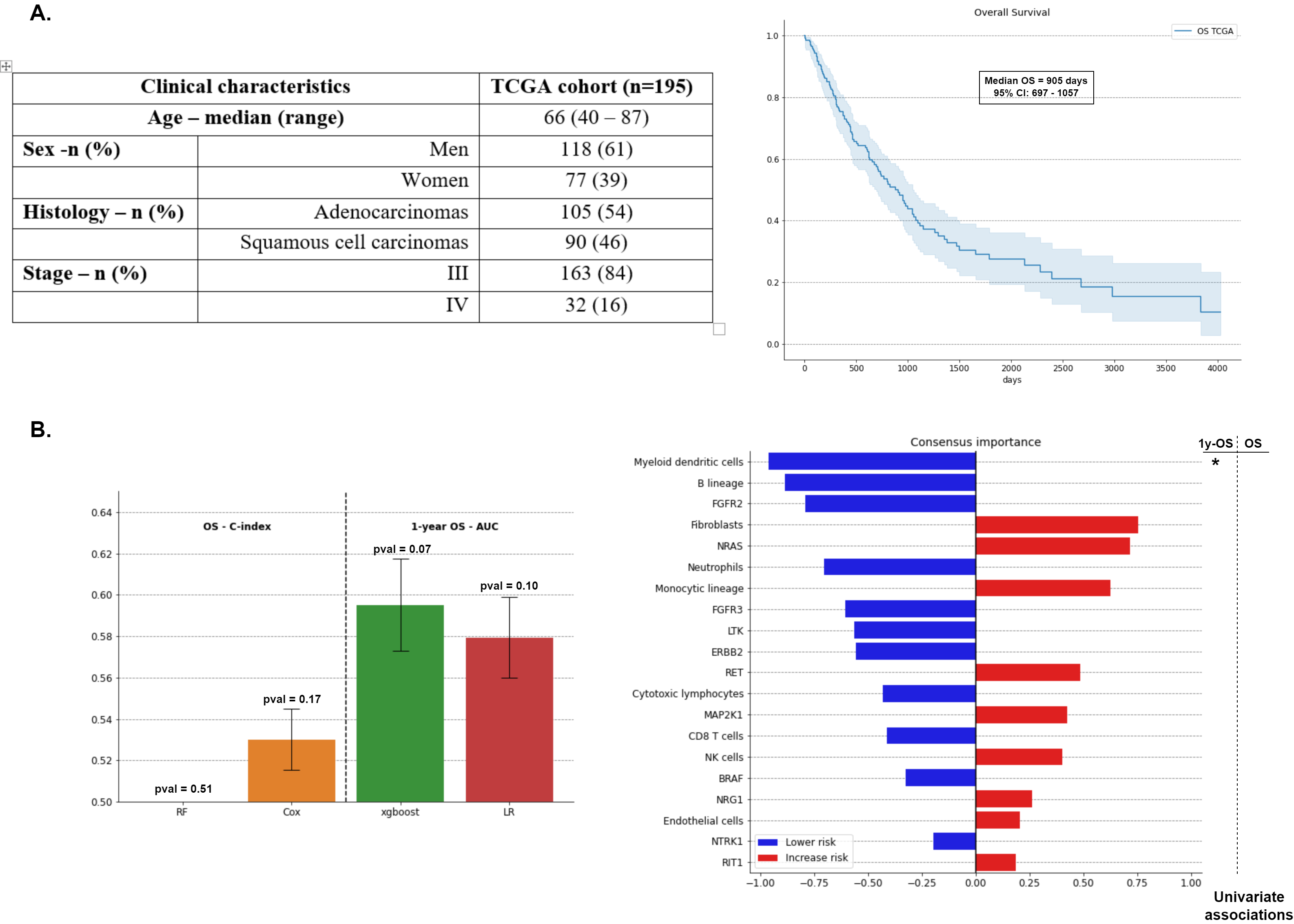

**Figure s24:** Prediction of OS and 1-year death for 195 stage III-IV NSCLC patients extracted from TCGA-LUSC and TCGA-LUAD. The exact same methodology as the one described in this analysis was used (i.e., 100 cross-validation schemes, XGBoost, Random Survival Forest, Logistic Regression, and Cox’s regression, and SHAP values for feature importance). **A.** Clinical characteristics of the TCGA cohort and OS Kaplan-Meier survival curve. **B.** Performance of the four predictive algorithms for the prediction of OS and 1-year death across the 100 cross-validation schemes (left). Feature importance ranking (i.e., aggregation of the SHAP values from both predictive tasks and both approaches) (right). The abundance of dendritic cells is the only feature that shows a significant association with 1-year death after Benjamini-Hochberg correction.

**Supplementary Tables**

**Table s1:** Permutation p-values for the unimodal performance of each data modality. P-values below 0.05 are indicated in red.

| **Target (number of patients)** | | **OS**  **(n=79)** | **1-year death**  **(n=77)** | **PFS**  **(n=80)** | **6-months progression**  **(n=75)** |
| --- | --- | --- | --- | --- | --- |
| **Metric** | | **C-index** | **AUC** | **C-index** | **AUC** |
| **Clinical** | Tree ensembles | < 0.01* | 0.07 | 0.07 | 0.06 |
|  | Linear | 0.03 | < 0.01* | 0.12 | 0.04 |
| **Radiomics** | Tree ensembles | 0.02 | 0.07 | 0.07 | 0.09 |
|  | Linear | 0.03 | 0.58 | 0.13 | 0.57 |
| **Pathomics** | Tree ensembles | 0.07 | 0.17 | 0.08 | 0.05 |
|  | Linear | 0.09 | 0.18 | 0.34 | 0.04 |
| **RNA** | Tree ensembles | <0.01* | < 0.01* | 0.06 | 0.05 |
|  | Linear | 0.07 | 0.06 | 0.03 | 0.09 |

*: p-values below 0.01 could not be estimated with a high precision since the number of permutations was limited to 100 due to computational constraints.
